# CD4^+^ T cell subset imbalances in rheumatoid arthritis

**DOI:** 10.1101/2022.10.20.22281307

**Authors:** B Mulhearn, L Marshall, M Sutcliffe, SK Hannes, C Fonseka, T Hussell, S Raychaudhuri, A Barton, S Viatte

**Affiliations:** Versus Arthritis Centre for Genetics and Genomics, Centre for Musculoskeletal Research, Manchester Academic Health Science Centre, The University of Manchester, Manchester, Oxford Road, Manchester, M13 9PT, UK; Lydia Becker Institute of Immunology and Inflammation, Faculty of Biology, Medicine and Health, The University of Manchester, Manchester, M13 9PL, UK; NIHR Manchester Musculoskeletal Biomedical Research Centre, Central Manchester NHS Foundation Trust, Manchester Academic Health Sciences Centre, Grafton Street, Manchester, M13 9WL, UK; Department of Life Sciences, University of Bath, Claverton Down, Bath, BA2 7AY, UK; Division of Rheumatology, Inflammation, and Immunity, Department of Medicine, Brigham and Women’s Hospital and Harvard Medical School, Boston, MA, USA; BioNTech, Cell Therapy Unit (Molecular Immunology), Cambridge, MA, USA; eGenesis, Cambridge, MA, USA

**Keywords:** precision medicine, rheumatoid arthritis, CD4^+^ T cells, mass cytometry, heterogeneity

## Abstract

**Background:** Despite the report of an imbalance between CD4^+^ T helper (Th) cell subsets in rheumatoid arthritis (RA), patient stratification for precision medicine has been hindered by the discovery of ever more Th cell subsets, as well as contradictory association results.

**Objectives:** To capture previously reported Th imbalance in RA with deep immunophenotyping techniques; to compare hypothesis-free unsupervised automated clustering with hypothesis-driven conventional biaxial gating and explore if Th cell heterogeneity accounts for conflicting association results.

**Methods:** Unstimulated and stimulated peripheral blood mononuclear cells from RA patients and controls were immunophenotyped with a 37-marker panel by mass cytometry (chemokine receptors, intra-cellular cytokines, intra-nuclear transcription factors). First, conventional biaxial gating and standard definitions of Th cell subsets were applied to compare subset frequencies between cases and controls. Second, unsupervised clustering was performed with FlowSOM and analysed using mixed-effects modelling of Associations of Single Cells (MASC).

**Results:** Conventional analytical techniques fail to identify classical Th subset imbalance, while unsupervised automated clustering, by allowing for unusual marker combinations, identified an imbalance between pro- and anti-inflammatory subsets. For example, a pro-inflammatory Th1-like (IL-2^+^ T-bet^+^) subset and an unconventional but pro-inflammatory IL-17^+^ T-bet^+^ subset were significantly enriched in RA (odds ratio=5.7, p=2.2 × 10^−3^; odds ratio=9.7, p=1.5×10^−3^, respectively). In contrast, a FoxP3^+^ IL-2^+^ HLA-DR^+^ Treg subset was reduced in RA (odds ratio=0.1, p=7.7×10^−7^).

**Conclusions:** Innovative approaches capture known CD4^+^ T cell subset imbalances in RA blood.

## Introduction

Current treatment strategies for rheumatoid arthritis (RA) standardise treatments across patient groups, but as a heterogeneous disease, differences will exist in the underlying immune mechanisms. As a consequence, not all patients will respond similarly to the same drug with only 60-70% having a response to any biologic drug [1]. Given that an increase of proinflammatory and/or a decrease of anti-inflammatory CD4^+^ T cell subsets has been reported in some patients, these cell types could represent therapeutic targets and aid patient stratification for precision medicine. However, which CD4^+^ T cell subset is associated with the disease, and in what proportion of patients, remains unclear.

The study of T cells in RA was revolutionised after the Th1/Th2 paradigm was proposed by Mossman et al. in 1986 [2]. This paradigm was described as a dichotomy between type 1 T helper cells (Th1), characterised by interferon-γ (IFN-γ) and tumour necrosis factor (TNF), and type 2 T helper cells (Th2). RA pathogenesis predominantly involves Th1 cytokines, and the Th1 immune response is antagonised by Th2 cytokines (reviewed in [3]). The Th1/Th2 paradigm was modified further when pathogenic IL-17-producing T helper cells (Th17) and regulatory T cells (Tregs) were discovered to play critical roles in initiating and regulating autoimmunity, respectively (reviewed in [4]). Increases in Th1 and Th17 cells which antagonise, and are antagonised by, Th2 and Treg cells has been the focus of much research in autoimmune diseases including RA [5,6]. More recently, interferon-producing Th1 memory cells were found to be associated with RA using novel techniques of analysing high-dimensional single-cell data [7]. A meta-analysis found that circulating Treg cells, as defined by both FOXP3 and CD25, were decreased in RA patients in 9 combined studies [8]. However, other studies found no decrease in Treg frequency or function in RA [9]. To add to the complexity of the paradigm, there are an ever-growing list of T helper cells (Th9 cells [10]; Th22 cells [11]; T follicular helper cells (Tfh) [12]; peripheral helper T cells (Tph) [13]). Critically, it has now been shown that many of these T cell types exhibit plasticity. It was previously thought that the mutually exclusive expression of a master transcription factor determined the fate of T cells, with T-bet, GATA3, RORγt and FoxP3 determining the fate of Th1, Th2, Th17 and Treg cells respectively. However, co-expression of these transcription factors may temporarily alter the effector function of T cell subsets (reviewed in [14]) and many of these T cells might also be transitional and therefore appear only in very low frequencies.

The plasticity of T cell subsets and heterogeneity of RA may also explain why clinical trials targeting specific cytokines, for instance IL-17, have not reached their primary endpoint [15]. Stratified medicine may allow clinicians to identify RA patients who have predominantly IL-17-driven disease to improve the design of such trials, as researchers have successfully shown in psoriatic arthritis [16]. Exploring peripheral blood immunophenotypes by flow cytometry in RA has been shown to mirror findings in the synovial compartment [17] and still represents the most widely used and cost effective technique to enumerate immune cell populations. The advent of mass cytometry (CyTOF) has vastly increased the number of cellular markers which can be detected simultaneously [18] and together with the development of high-throughput automated methods of data analysis [19–23], rare and novel cell types involved in the aetiology of RA have been recently identified [7,13,24,25]. Automated cell clustering algorithms allow for unbiased marker combinations and therefore the hypothesis-free discovery of unanticipated cell subsets, relevant to disease and precision medicine [7,22,26,27]. We postulate that Th cell plasticity and overlap between subsets and definitions explain conflicting associations and lack of reproducibility in small sample sizes. Therefore, we developed a 37 marker T cell mass cytometry panel to encompass most definitions for the most studied CD4^+^ T cell subsets to date (Th1, Th2, Th17 and Treg).

The aims of this study are first, to confirm the T helper cell subset imbalance previously described in RA; second, to explore whether this imbalance is detectable in a small sample size, if the standard definition of Th subsets is relaxed in favour of Th cell plasticity (unbiased marker combinations) and third, to test whether innovative techniques (mass cytometry and unsupervised clustering algorithm) facilitate the identification of pro- and anti-inflammatory CD4^+^ T cell subsets over standard techniques (flow cytometry and manual bi-axial gating) in a small sample size.

## Methods

### Patient and public involvement

The Versus Arthritis Centre for Genetics and Genomics in Manchester has a Research User Group (RUG) of patients with various rheumatologic conditions, which includes RA. Members of the RUG meet regularly to review the research carried out in the Centre. They highlighted the importance of understanding basic mechanisms of disease from a patient’s perspective. The RUG supports research of basic disease mechanisms to identify biomarkers for stratification into treatment response categories. Some of the following comments were made: “the sooner anyone with RA can get on the correct treatment, the better”; “Personalising from onset would be perfect”; “If only a blood test is needed, then even better”.

### Patient selection and sample collection

A cohort of 10 RA patients stabilised on therapy and 10 healthy volunteers were recruited from the National Repository (North West Ethics committee approval MREC 99/8/84) at the Manchester Royal Infirmary (NIHR portfolio ID 7881). Peripheral Blood Mononuclear Cells (PBMC) were isolated from 18 ml of blood by density gradient centrifugation using Ficoll-Paque plus (GE Healthcare Life Sciences) and cryopreserved at -150°C.

### Mass cytometry antibody panel

We adapted a previously published mass cytometry T cell panel [13] to encompass surface (chemokine receptors), intra-cytoplasmic (cytokines) and intra-nuclear markers (transcription factors) used to define Th and Treg subsets (Table 1)

**Table 1.**
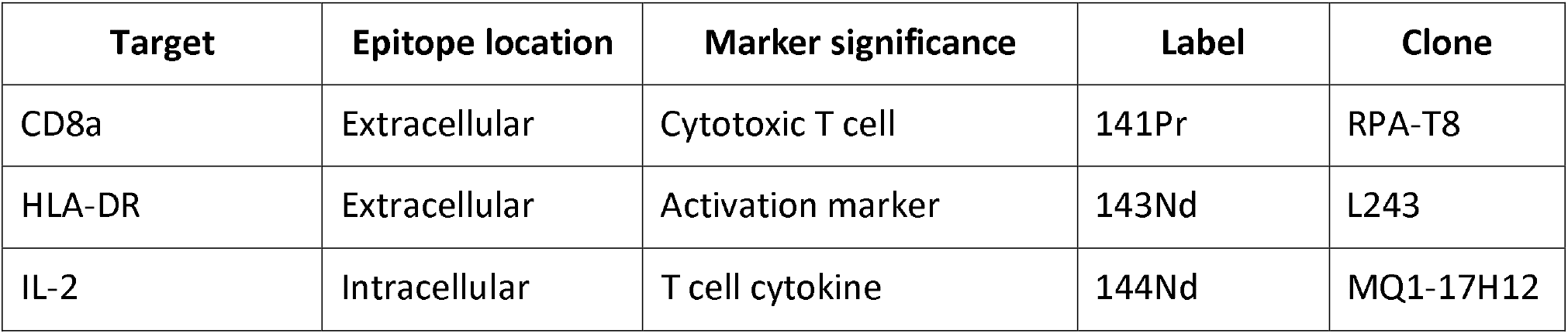
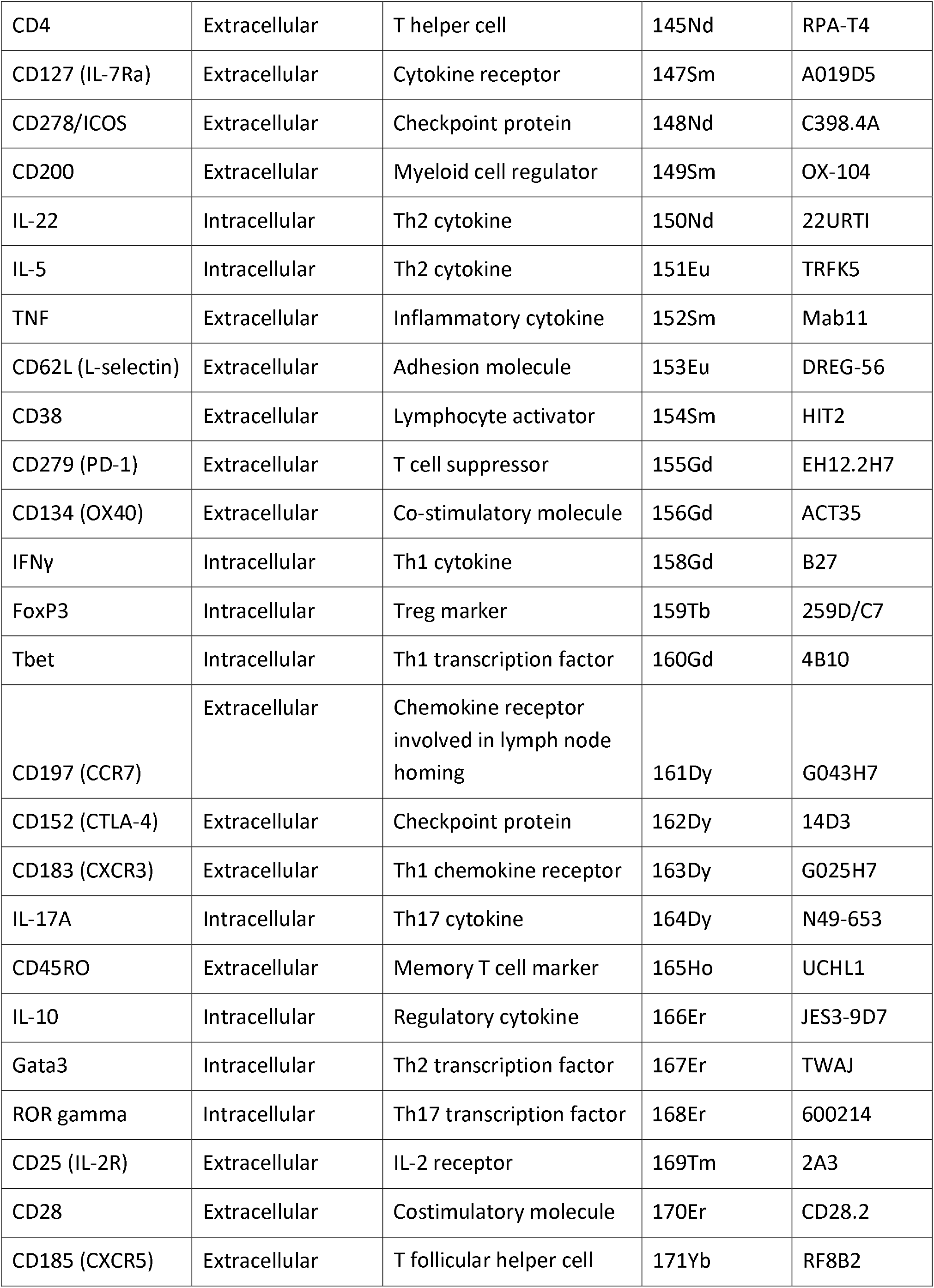
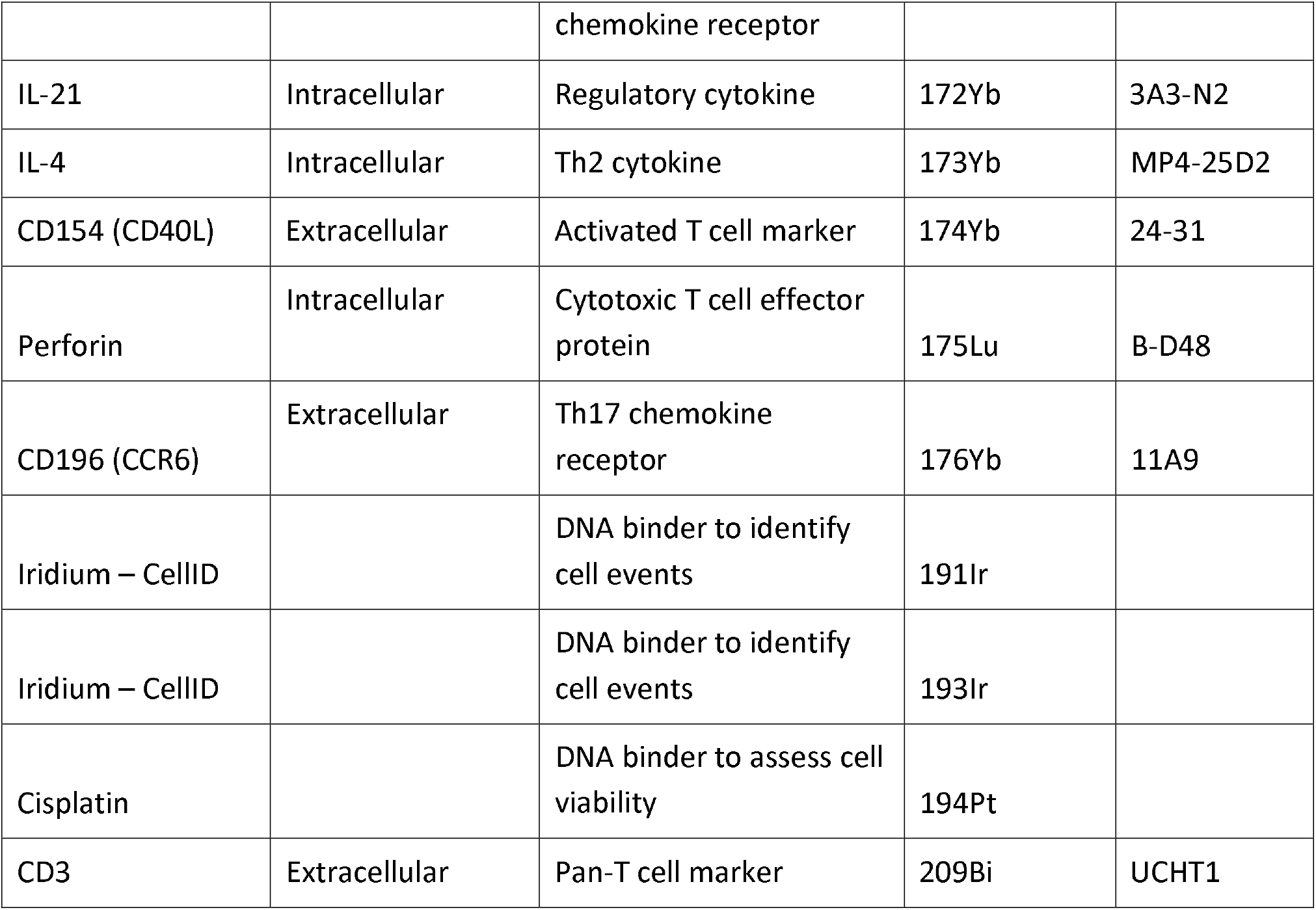
The antigen target, heavy metal and clone of each conjugated antibody used in the CyTOF T cell panel. Target location indicates if the antibody was used in the extracellular or intracellular antibody cocktail. All antibodies were purchased from Fluidigm.

### T cell enrichment and stimulation

Thawed PBMCs were rested at 37°C for one hour prior to enrichment of CD3^+^ T cells by positive selection using magnetic cell separation (MACS) with CD3 MicroBeads (Miltenyi Biotec). T cell receptor stimulation was achieved with Dynabeads Human T-Activator CD3/CD28 beads (Fisher Scientific UK Ltd) at a concentration of 1 bead per 2 cells, in the presence of brefeldin A and monensin (both Fisher Scientific UK Ltd) and incubated for 4.5 hours at 37 °C.

### Mass cytometry staining protocol

CD3^+^ T cells were incubated with cisplatin prior to incubation with an extracellular antibody cocktail (detailed protocol in Supplementary Methods). Subsequently, cells were fixed, permeabilised and stained for intracellular antigens for 30 minutes on ice. A solution of Intercalator-Ir was added to each well. Samples were run on Helios mass cytometers by the Longwood Medical Area CyTOF Core Facility at the Dana-Farber Cancer Institute, Boston, USA.

### Data pre-processing

The Nolan Normalizer MATLAB plugin [28] was used to normalise the signal in each channel to the signals from the Maxpar Calibration beads.

### Traditional gating analysis

Traditional biaxial gating was carried out using FlowJo V.10.8 (BD Biosciences) to identify well-described CD4^+^ Th and Treg subsets using standard definitions [29] (example gating strategy in supplementary Fig. 1). The percentages of positive cells are represented as a proportion of the total T cell population (CD3^+^). Geometric mean intensity is used to quantify cytokine expression on an individual cell level. The Mann-Whitney U test was used to compare RA and Healthy Controls (HC). All p-values are unadjusted (tests are not independent).

### Automated clustering workflow

The CyTOF clustering workflow from Nowicka et al. [20] was modified to include an extended quality control approach and a different statistical framework for association testing (see below). All plots were produced with *ggplot2* (v3.3.5) [30], unless stated otherwise.

#### Quality control steps

Normalised cytometric data was manually inspected using FlowJo V.10.8 (BD Biosciences) to ensure that at least 10 cell events were identified in a cluster in at least 3 samples. IL-5 was excluded due to the absence of any positive cell clusters (supplementary figure 2). Furthermore, only samples with at least 1000 live single T cells were included in subsequent analyses. Cell events with extreme expression were excluded per individual marker. After the removal of unsuccessful markers and extreme events, analysis was performed in R (v4.1.0) on data transformed with the inverse hyperbolic sine (arcsinh) function (cofactor = 5). Finally, to identify outlying samples and potential batch effects, multidimensional scaling (MDS) and principal component analysis (PCA) plots were produced from median expression of panel markers in each sample.

#### Clustering with FlowSOM/ConsensusClusterPlus

The dataset was downsampled to an equal number of randomly selected cells (n) from each sample, where n was equal to the number of cells in the smallest sample. The automated clustering steps were performed with *FlowSOM* (v2.0.0) [19] and *ConsensusClusterPlus* (v1.56.0) [23] using agglomerative hierarchical consensus clustering. Cells were clustered into 20 populations using Euclidean distance and average linkage metrics (supplementary figure 3). Heatmaps of median marker expression for each cluster were generated with *pheatmap* (v1.0.12) [31]. For visualisation, these values were scaled between 0 and 1, with the 0.01 and 0.99 quantiles for each marker as the lower and upper boundary, respectively.

#### Cluster visualisation with t-SNE

The similarity of single cells in two-dimensional space was visualised with the dimensionality reduction technique t-stochastic neighbour embedding (t-SNE), implemented with *Rtsne* (v0.15) [32]. Clusters were annotated manually by their surface phenotype.

### Statistical analysis

To identify clusters with differential representation in RA samples compared to healthy controls, we used mixed-effects modelling of associations of single cells (MASC) [7] for each cluster, including age and sex of donors as fixed-effects covariates, sample ID as a random-effect covariate and the case-control status as the contrast term. Clusters that are significantly enriched or depleted in RA are defined as those with a P value ≤ 0.05 after Bonferroni correction (n = number of clusters tested).

### Data sharing

All mass cytometry data is freely available at https://flowrepository.org/ (repository ID FR-FCM-Z5RM) [46].

## Results

### Study design and subject characteristics

PBMCs from 10 RA patients and 10 healthy controls (Table 2) were left unstimulated or stimulated with anti-CD3/CD28 beads (*n* = 40 samples) prior to deep immunophenotyping by mass cytometry.

**Table 2.** Subject characteristics of cases (RA) and healthy controls. *P* values calculated for age and gender by Mann-Whitney U and Fisher’s exact test, respectively. RF: rheumatoid factor. ACPA: anti-citrullinated protein antibody. DAS: disease activity score. DMARD: disease-modifying anti-rheumatic drug.

| | Cases ($n = 10$) | Controls ($n = 10$) | *P* value |
| --- | --- | --- | --- |
| **Age (mean ± SD)** | 61 ± 14 | 46 ± 8 | 0.02 |
| **Female** | 7 | 6 | 1 |
| **RF-positive** | 7 | — | — |
| **ACPA-positive** | 6 | — | — |
| **DAS28 (mean ± SD)** | 3.4 ± 2.2 | — | — |
| **Glucocorticoids** | 2 | 0 | — |
| **Methotrexate** | 5 | 0 | — |
| **Other DMARD\*** | 3 | 0 | — |
| **Biologics** | 1 | 0 | — |

\*Sulfasalazine or hydroxychloroquine

*Sulfasalazine or hydroxychloroquine

### Manual gating with traditional statistical analysis fails to identify large differences between blood from patients with RA and healthy controls

Sequential biaxial gating of CyTOF data was used to identify commonly described CD4^+^ and CD8^+^ T cell subsets as defined by expression of surface markers, transcription factors and/or intracellular cytokines. Conventional statistical analysis with Mann-Whitney *U* found no significant differences in the proportions of Th CD4^+^ subsets between RA and HC (figure 1); only Treg (defined as CD4^+^ FoxP3^+^) were markedly decreased in RA. No CD8^+^ T cell subsets were found to be differentially abundant in RA (figure 2).

**Figure 1.**
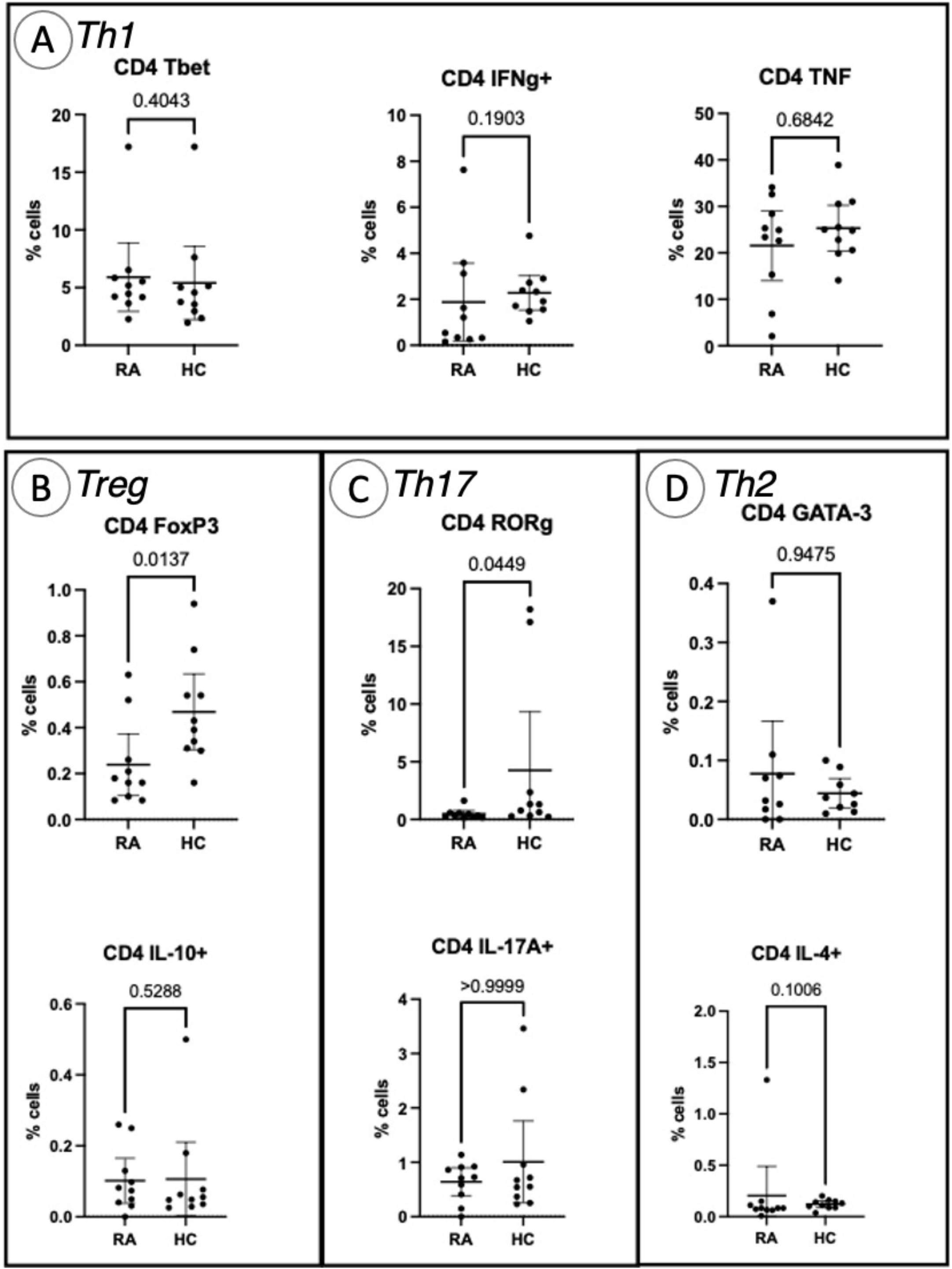
Abundances of Th and Treg CD4^+^ T cell subsets identified by manual gating of CyTOF data shown with mean, 95% confidence interval and p-value (shown above the pairwise comparisons). Points represent RA and HC samples. Mann Whitney *U* P values are shown above the pairwise comparisons. Graphs with cytokine expression are taken from the stimulated T cell dataset and those with surface markers and transcription factors are taken from the unstimulated dataset. Box A represents markers associated with Th1 cells, box B Treg cells, box C represents Th17-related markers, and box D Th2-related markers.

**Figure 2.**
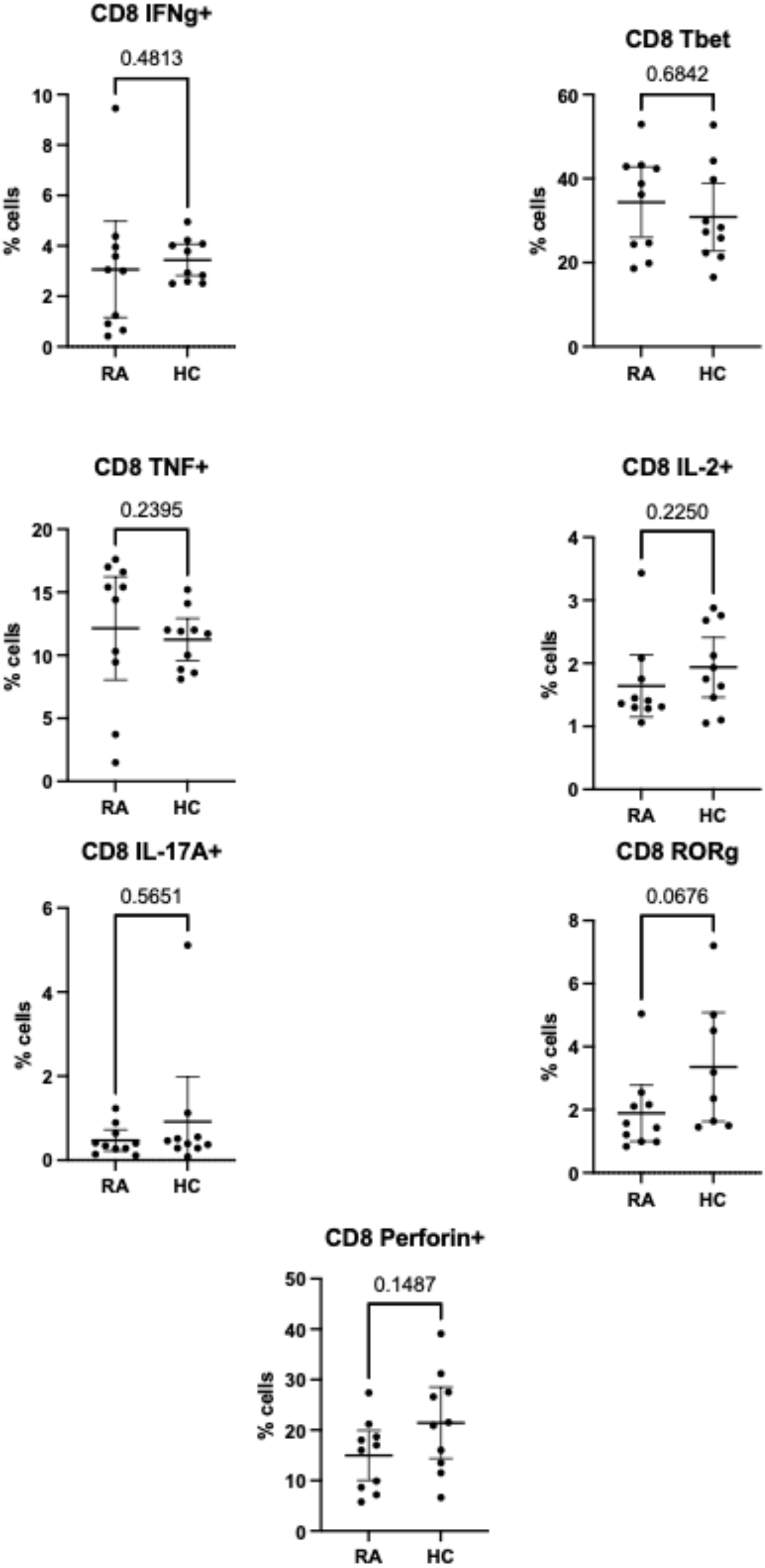
Abundances of CD8^+^ T cell subsets identified by manual gating of CyTOF data shown with mean and 95% confidence interval. Points represent RA and HC samples. Mann Whitney *U* P values are shown above the pairwise comparisons. Graphs with cytokine expression are taken from the stimulated T cell dataset and those with surface markers and transcription factors are taken from the unstimulated dataset.

### CyTOF automated clustering pipeline quality control steps

Low quality cells with extreme expression values were excluded after individual appraisal of expression distributions for each marker. This step removed 0.34% (4,146/1,230,364) of cells from the full dataset, with between 0.09% and 1.4% of cells removed from each sample. Diagnostic MDS and PCA plots highlight notable separation of RA and HC in the second dimension, indicating preliminary differences in marker expression between the two groups (figure 3A). On the same PCA (figure 3B), samples largely cluster by age group (figure 3C) similarly to previous reports [49], suggesting that the structure in the data is attributable to participant age rather than technical batch effects. Therefore, we included age as a co-variate for statistical association testing.

**Figure 3.**
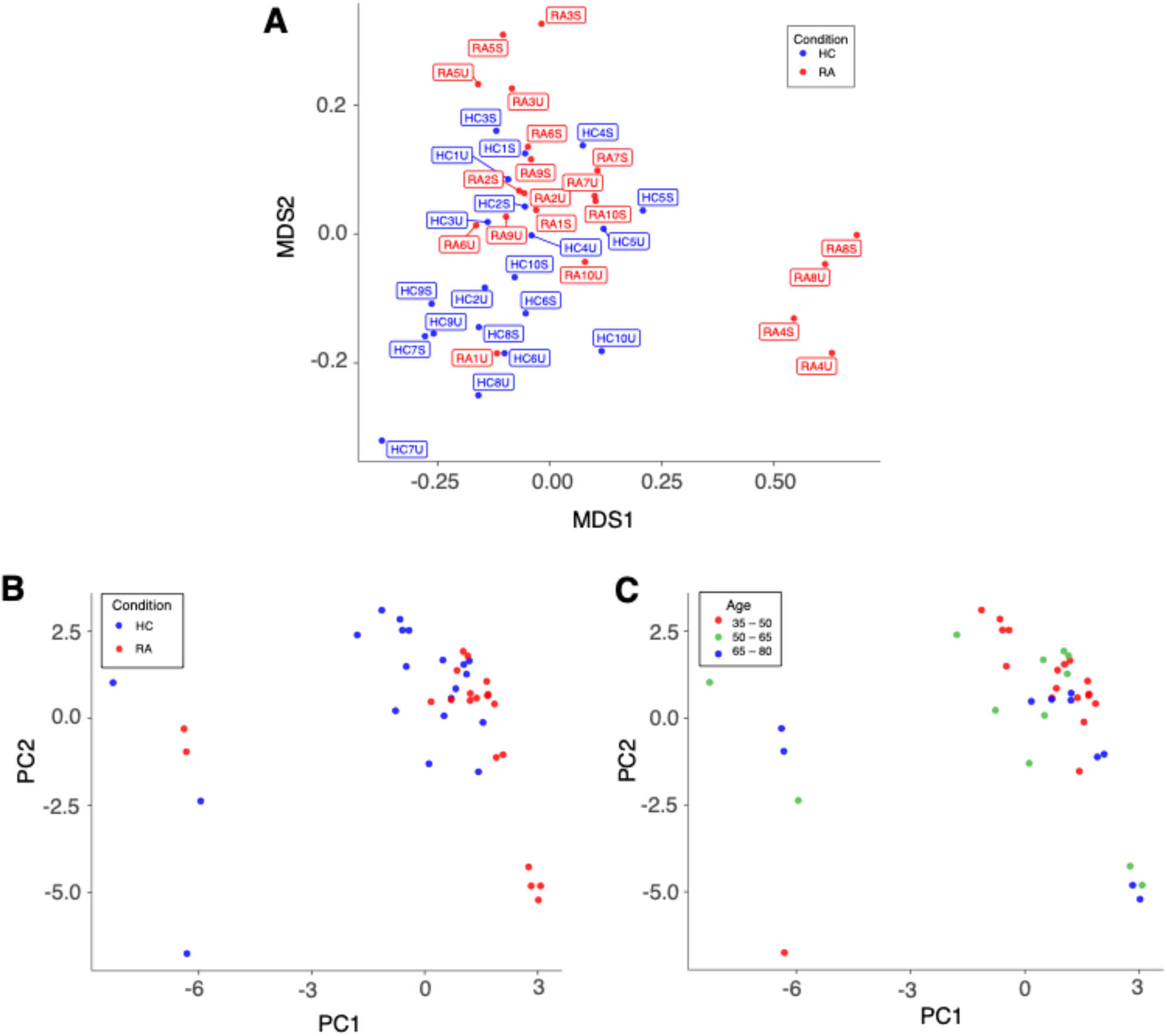
**A)** Multidimensional scaling (MDS) plot based on median, arcsinh-transformed marker expression in unstimulated (U) and stimulated (S) T cell samples from rheumatoid arthritis (RA) patients and healthy controls (HC). **B**) PCA plot of the same data coloured by case-control status. **C)** PCA plot of the same data coloured by age group of participants.

### Automated clustering identifies well-described T cell subsets

Automated clustering was performed on downsampled quality-controlled CyTOF data; analyses of unstimulated and stimulated T cell datasets were performed separately. Within the unstimulated dataset, we identified 20 distinct T cell subsets, including 11 CD4^+^ and nine CD8^+^ (figure 4A). Amongst the CD4^+^ populations are a FoxP3^+^ subset (subset 11; a Treg subset), two IL-17^+^ subsets (subsets 16 and 20; Th17 subsets), a Th1-like subset (subset 19) and a CD38^+^ subset (subset 18).

**Figure 4.**
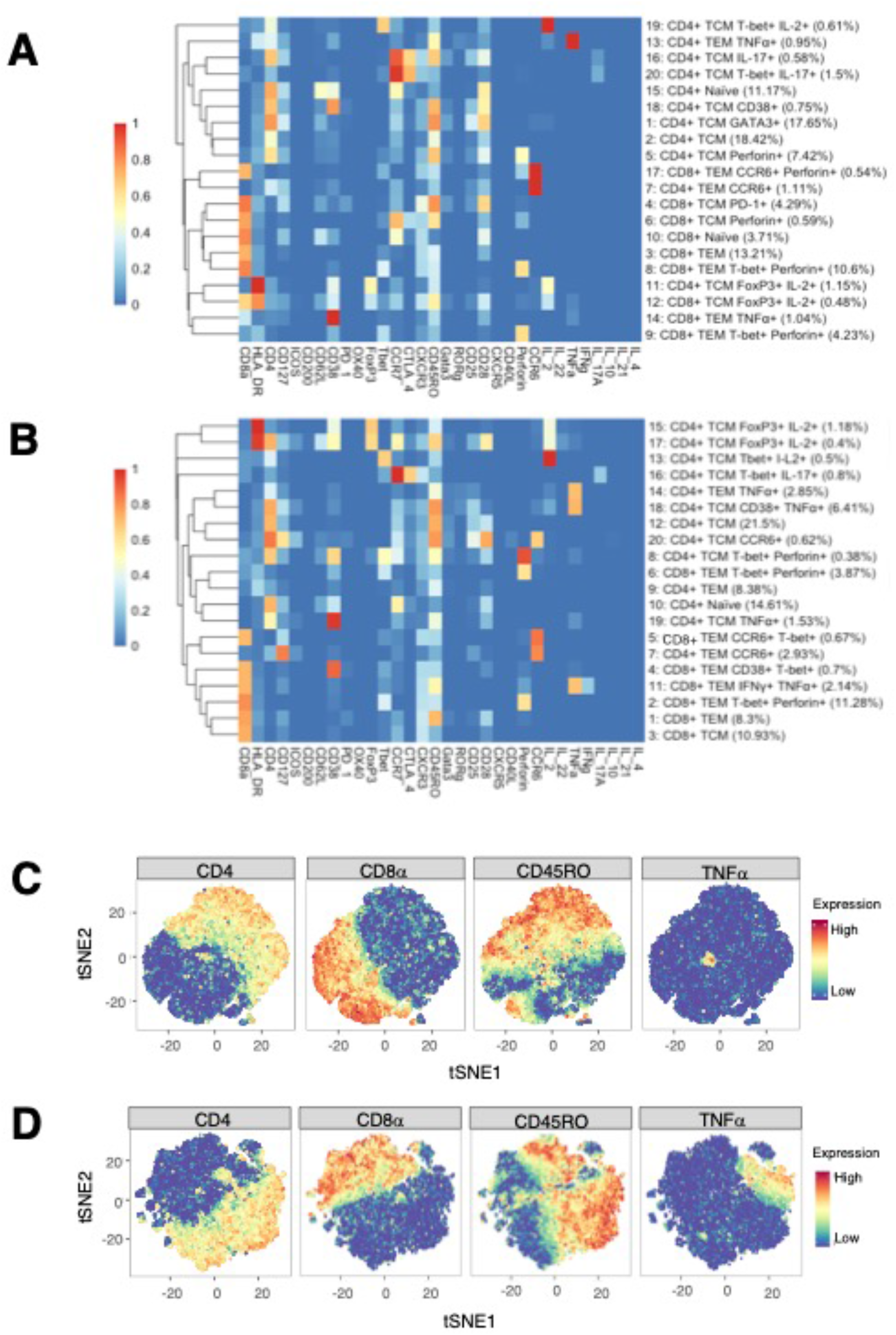
**A)** Heatmap of median marker expression (arcsinh, 0-1 transformed) for 20 T cell clusters identified in unstimulated cell samples by automated clustering. Each sample was downsampled to 2746 cells (n = 10 RA, 10 HC). The dendrogram represents hierarchical clustering using Euclidean distance and average linkage. **B)** As A), for stimulated samples (*n* = 10 RA, 10 HC). Each sample was downsampled to 2224 cells. C) t-SNE plots of the same unstimulated cells, coloured by expression level of a selection of markers. D) As C), for stimulated cells.

Although analysis was performed independently, clusters identified in the stimulated dataset were broadly similar to those of the unstimulated dataset (figure 4B). As expected, t-SNE plots coloured by expression level of several markers highlight an increase in the number of cells expressing TNF upon stimulation (figures 4B and 4C).

### MASC identifies multiple T cell clusters with differential expression between RA and healthy samples

MASC was used to identify clusters with differential abundance between RA samples and HC, adjusting for the effects of age, gender, and donor. Six unstimulated cell subsets and 8 stimulated subsets were found to be differentially abundant with MASC (figure 5). The majority were CD4^+^ and, of these, three populations were present in both unstimulated and stimulated states, including enrichment of Th1 and Th17 subsets (CD4^+^ T-bet^+^ IL-17^+^ and CD4^+^ T-bet^+^ IL-2^+^ cell clusters) and depletion of a Treg subset (CD4^+^ FoxP3^+^ IL-2^+^) in RA (figure 6). Of note, stimulation further enriched the CD4^+^ T-bet^+^ IL-2^+^ cluster in RA samples relative to healthy controls (P = 2.2 × 10^−3^, OR = 5.7, figure 7). Interestingly, a Th2 subset (CD4^+^ T_CM_ GATA3^+^) was decreased in RA (although it did not reach statistical significance).

**Figure 5.**
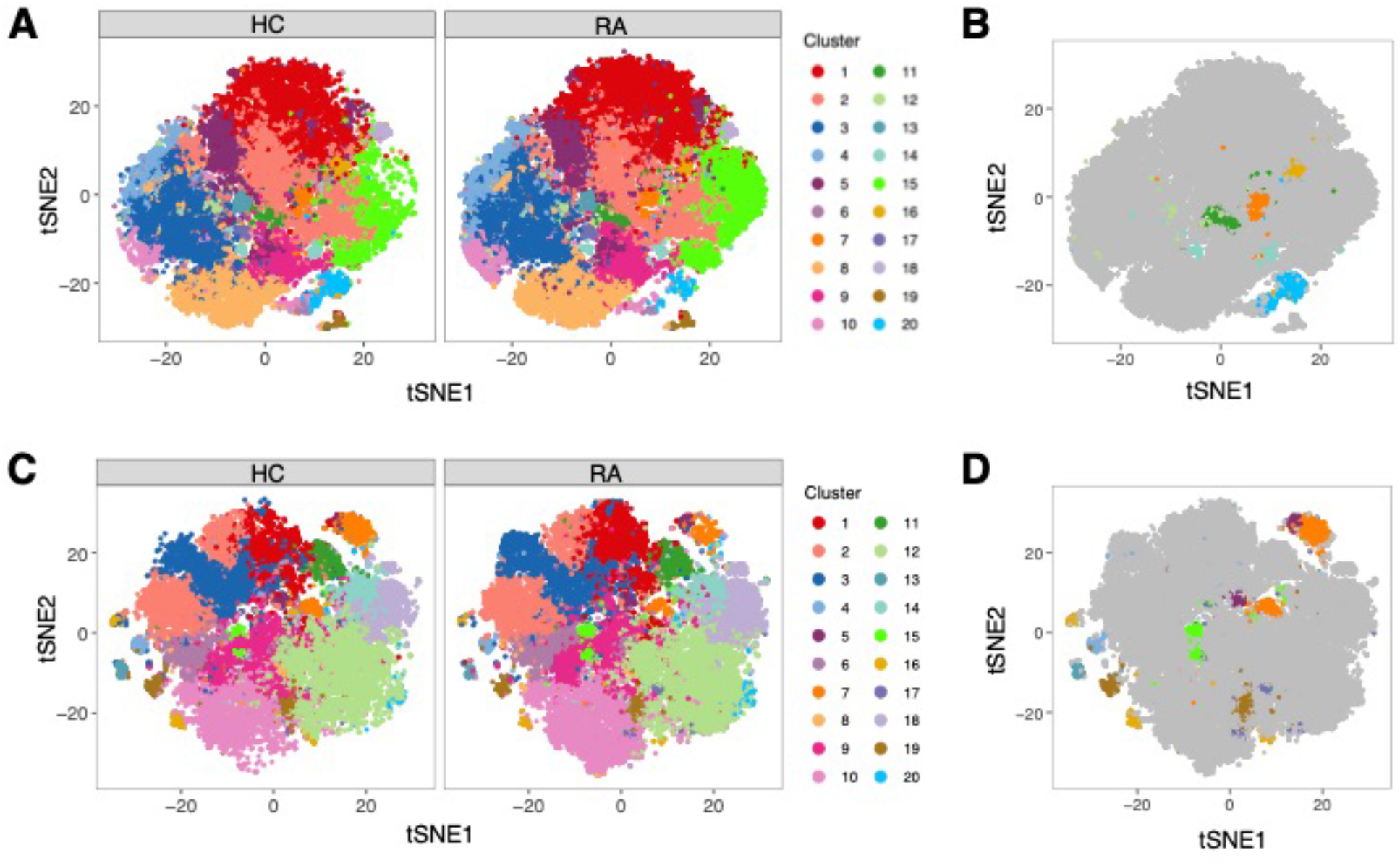
**A)** tSNE plots of unstimulated cells faceted by case-control status and coloured by cluster assignment. Samples were downsampled to 2746 cells each (*n* = 20). **B)** tSNE plot of unstimulated cells with conditions combined. Clusters with differential abundance in RA vs healthy controls (HC) according to MASC are highlighted (*P* ≤ 0.05). **C**) As A), for stimulated cells. Samples were downsampled to 2224 cells each (*n* = 20). **D)** As B), for unstimulated cells. Note that unstimulated and stimulated samples were clustered independently.

**Figure 6.**
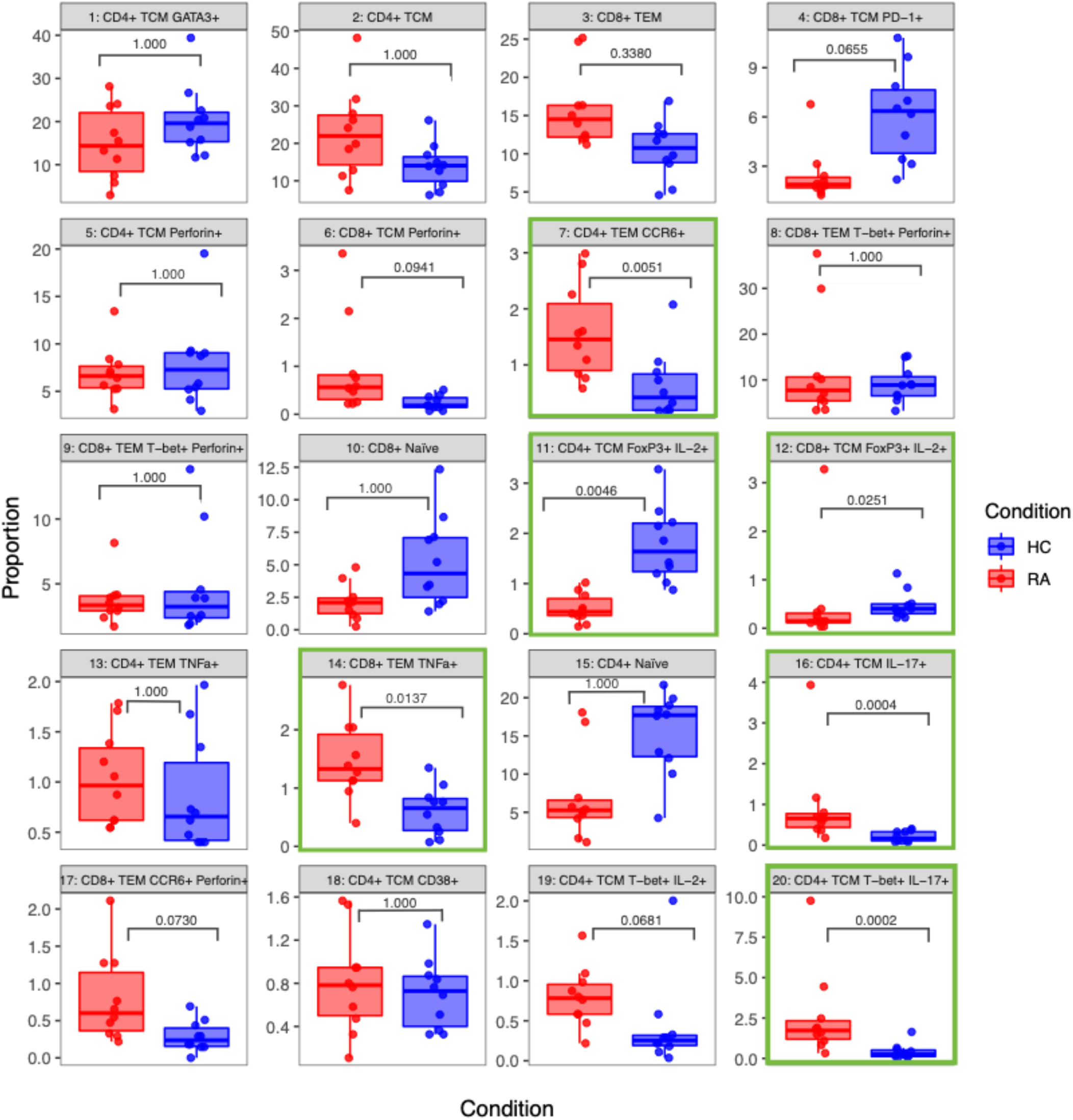
Boxplots of the proportions of 20 T cell clusters identified in unstimulated RA and healthy control (HC) samples (*n* = 10 RA, 10 HC). The P value obtained from MASC is shown for each cluster, and a green outline indicates differential abundance between RA and HC (*P* ≤ 0.05).

**Figure 7.**
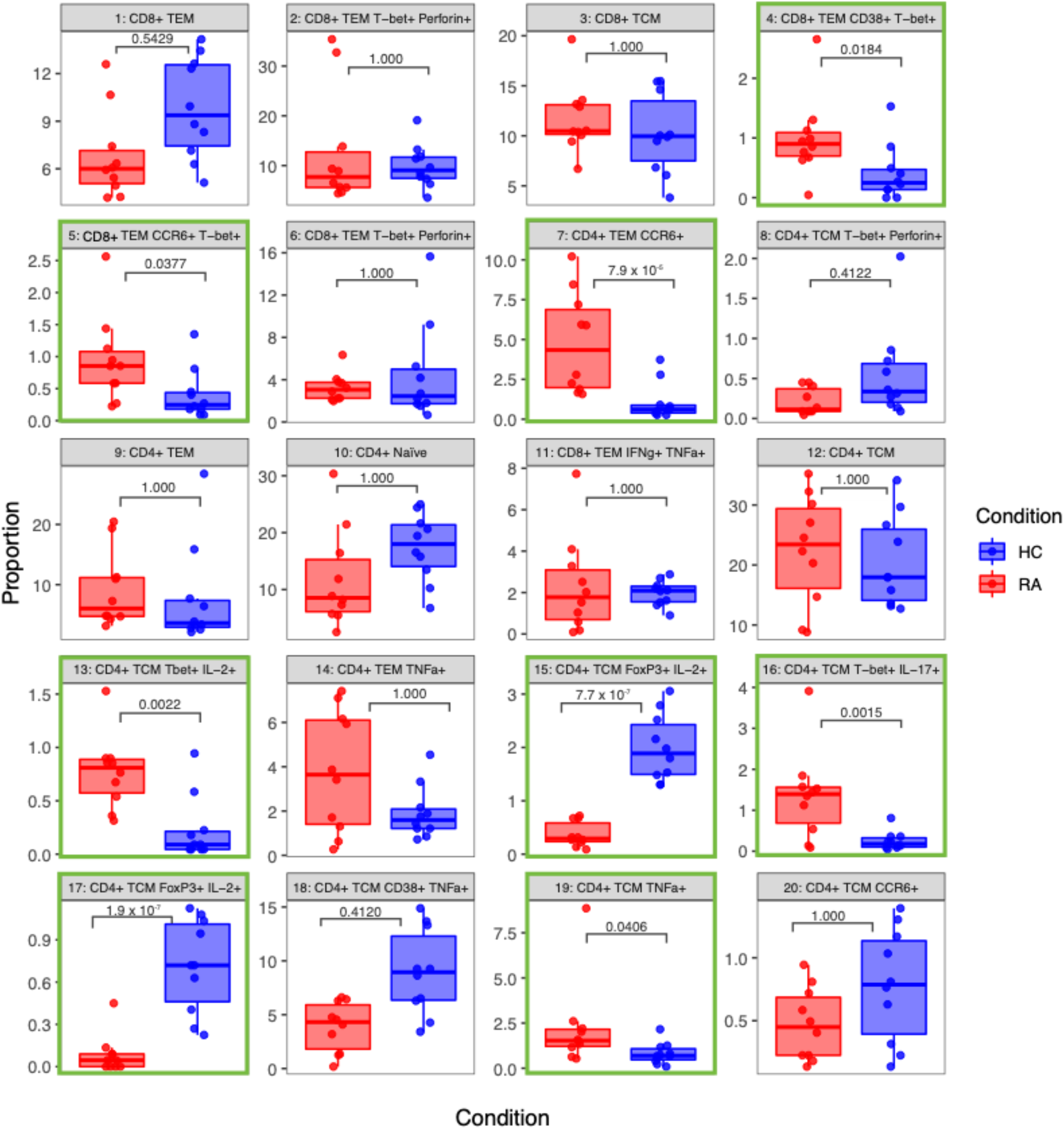
Boxplots of the proportions of 20 T cell clusters identified in stimulated RA and healthy control (HC) samples (*n* = 10 RA, 10 HC). The *P* value obtained from MASC is shown for each cluster, and a green outline indicates differential abundance between RA and HC (*P* ≤ 0.05).

Notably, figures 6 and 7 highlight the heterogeneity in subset proportions between individual patients. For example, the abundance of the CD4^+^ TNF^+^ cells range from < 1% to > 8% of cells in stimulated RA samples (figure 7, subset 14). Naïve T cells were increased in frequency in controls compared to cases, but this difference disappeared after statistical adjustment for age implemented in MASC, demonstrating that the correction for the age-driven stratification observed in the PCA plots was important in terms of statistical outcome.

Cell cluster phenotype, abundances, odds ratios, and P values for all cell clusters generated by the pipeline are presented in table 3.

**Table 3.** MASC results for clusters present in both unstimulated and stimulated samples (A), unstimulated samples alone (B) and stimulated samples alone (C). For shared cell subsets, the first row indicates statistics for unstimulated subsets and the second for stimulated subsets. Elements separated by “/” indicates statistics for two identical subsets found in the unstimulated and stimulated datasets. The odds ratio is shown with a 95% confidence interval. Differentially abundant subsets are highlighted by a bold P value (α = 0.05). T_CM_: T central memory. T_EM_: T effector memory.

| Subset | Number | % total<br>RA cells | % total<br>HC cells | OR (RA vs HC) | P value |
| --- | --- | --- | --- | --- | --- |
| **A: Shared (Unstimulated, stimulated)** |  |  |  |  |  |
| **CD4<sup>+</sup> T<sub>CM</sub> T-bet<sup>+</sup> IL-17<sup>+</sup>**<br>(combined Th1 and Th17 phenotype) | 20 | 2.6 | 0.4 | 12.2 (5.3 – 28.1) | **1.5 x 10<sup>-4</sup>** |
|  | 16 | 1.4 | 0.2 | 9.7 (3.8 – 24.5) | **1.5 x 10<sup>-3</sup>** |
| **CD4<sup>+</sup> T<sub>CM</sub> FoxP3<sup>+</sup> IL-2<sup>+</sup>**<br>(Treg) | 11 | 0.5 | 1.8 | 0.3 (0.2 – 0.5) | **4.6 x 10<sup>-3</sup>** |
|  | 15 + 17 | 0.4 / 0.09 | 2.0 / 0.7 | 0.1 (0.1 – 0.2) /<br>0.03 (0.01 – 0.09) | **7.7 x 10<sup>-7</sup> /**<br>**1.9 x 10<sup>-7</sup>** |
| **CD4<sup>+</sup> T<sub>EM</sub> CCR6<sup>+</sup>**<br>(Th17) | 7 | 1.6 | 0.6 | 4.4 (2.2 – 8.7) | **5.1 x 10<sup>-3</sup>** |
|  | 7 | 4.8 | 1.1 | 9.4 (4.6 – 19.5) | **7.9 x 10<sup>-5</sup>** |
| **CD4<sup>+</sup> T<sub>CM</sub> T-bet<sup>+</sup> IL-2<sup>+</sup>**<br>(Th1) | 19 | 0.8 | 0.4 | 3.5 (1.6 – 7.4) | 6.8 x 10<sup>-2</sup> |
|  | 13 | 0.8 | 0.2 | 5.7 (2.6 – 12.3) | **2.2 x 10<sup>-3</sup>** |
| **CD8<sup>+</sup> T<sub>EM</sub>** | 3 | 15.9 | 10.5 | 1.7 (1.1 – 2.7) | 3.4 x 10<sup>-1</sup> |
|  | 1 | 6.8 | 9.8 | 0.6 (0.4 – 0.9) | 5.4 x 10<sup>-1</sup> |
| **CD4<sup>+</sup> T<sub>CM</sub>** | 2 | 22.8 | 14.0 | 1.5 (0.9 – 2.7) | 1 |
|  | 12 | 22.5 | 20.6 | 0.9 (0.5 – 1.5) | 1 |
| **CD8<sup>+</sup> T<sub>EM</sub> T-bet<sup>+</sup> Perforin<sup>+</sup>** | 8 + 9 | 12.2 / 3.7 | 9.0 / 4.8 | 1.0 (0.5 – 1.9) /<br>0.8 (0.4 – 1.5) | 1 |
|  | 2 | 12.7 | 9.9 | 0.9 (0.5 – 1.9) | 1 |
| **CD4<sup>+</sup> T<sub>EM</sub> TNFα<sup>+</sup>**<br>(Th1) | 13 | 1.0 | 0.9 | 1.1 (0.6 – 1.9) | 1 |
|  | 14 | 3.8 | 1.9 | 1.9 (0.8 – 4.7) | 1 |
| **CD4<sup>+</sup> Naïve** | 15 | 7.0 | 15.4 | 0.6 (0.3 – 1.1) | 1 |
|  | 10 | 11.8 | 17.4 | 0.8 (0.4 – 1.4) | 1 |
| **B: Unstimulated** |  |  |  |  |  |
| **CD4<sup>+</sup> T<sub>CM</sub> IL-17<sup>+</sup>**<br>(Th17) | 16 | 0.9 | 0.2 | 5.7 (3.1 – 10.3) | **4.0 x 10<sup>-4</sup>** |
| **CD8<sup>+</sup> T<sub>EM</sub> TNFα<sup>+</sup>** | 14 | 1.5 | 0.6 | 3.7 (2.0 – 7.1) | **1.4 x 10<sup>-2</sup>** |
| **CD8<sup>+</sup> T<sub>CM</sub> FoxP3<sup>+</sup> IL-2<sup>+</sup>** | 12 | 0.5 | 0.5 | 0.2 (0.1 – 0.5) | **2.5 x 10<sup>-2</sup>** |
| **CD8<sup>+</sup> T<sub>CM</sub> PD-1<sup>+</sup>** | 4 | 2.4 | 6.2 | 0.4 (0.2 – 0.7) | 6.5 x 10<sup>-2</sup> |
| **CD8<sup>+</sup> T<sub>EM</sub> CCR6<sup>+</sup> Perforin<sup>+</sup>** | 17 | 0.8 | 0.3 | 3.5 (1.6 – 7.6) | 7.3 x 10<sup>-2</sup> |
| **CD8<sup>+</sup> T<sub>CM</sub> Perforin<sup>+</sup>** | 6 | 0.9 | 0.2 | 3.8 (1.6 – 8.7) | 9.4 x 10<sup>-2</sup> |
| **CD4<sup>+</sup> T<sub>CM</sub> GATA3<sup>+</sup>** | 1 | 15.0 | 20.3 | 0.6 (0.3 – 1.2) | 1 |
| **CD4<sup>+</sup> T<sub>CM</sub> Perforin<sup>+</sup>** | 5 | 6.9 | 7.9 | 0.7 (0.4 – 1.2) | 1 |
| **CD8<sup>+</sup> Naïve** | 10 | 2.2 | 5.3 | 0.8 (0.4 – 1.5) | 1 |
| **CD4<sup>+</sup> T<sub>CM</sub> CD38<sup>+</sup>** | 18 | 0.8 | 0.7 | 1.6 (1.0 – 2.7) | 1 |
| **C: Stimulated** |  |  |  |  |  |
| **CD8<sup>+</sup> T<sub>EM</sub> CD38<sup>+</sup> T-bet<sup>+</sup>** | 4 | 1.0 | 0.4 | 6.3 (2.3 – 17.0) | **1.8 x 10<sup>-2</sup>** |
| **CD8<sup>+</sup> T<sub>EM</sub> CCR6<sup>+</sup> T-bet<sup>+</sup>** | 5 | 0.9 | 0.4 | 3.8 (1.8 – 7.9) | **3.8 x 10<sup>-2</sup>** |
| **CD4<sup>+</sup> T<sub>CM</sub> TNFα<sup>+</sup>** | 19 | 2.2 | 0.8 | 4.0 (1.9 – 8.5) | **4.1 x 10<sup>-2</sup>** |
| **CD4<sup>+</sup> T<sub>CM</sub> CD38<sup>+</sup> TNFα<sup>+</sup>** | 18 | 3.9 | 9.0 | 0.3 (0.1 – 0.8) | 4.1 x 10<sup>-1</sup> |
| **CD4<sup>+</sup> T<sub>CM</sub> T-bet<sup>+</sup> Perforin<sup>+</sup>** | 8 | 0.2 | 0.6 | 0.3 (0.1 – 0.8) | 4.1 x 10<sup>-1</sup> |
| **CD8<sup>+</sup> T<sub>CM</sub>** | 3 | 11.7 | 10.2 | 1.4 (0.9 – 2.0) | 1 |
| **CD8<sup>+</sup> T<sub>EM</sub> T-bet<sup>+</sup> Perforin<sup>+</sup>** | 6 | 3.3 | 4.5 | 0.9 (0.4 – 2.0) | 1 |
| **CD4<sup>+</sup> T<sub>EM</sub>** | 9 | 9.1 | 7.6 | 1.1 (0.5 – 2.4) | 1 |
| **CD8<sup>+</sup> T<sub>EM</sub> IFNγ<sup>+</sup> TNFα<sup>+</sup>** | 11 | 2.3 | 2.0 | 0.9 (0.4 – 2.3) | 1 |
| **CD4<sup>+</sup> T<sub>CM</sub> CCR6<sup>+</sup>** | 20 | 0.5 | 0.8 | 0.7 (0.4 – 1.5) | 1 |

### Agnostically defined cell clusters can be identified using conventional techniques

To provide internal validation of the automated pipeline’s significant clusters, manual gating and standard statistical analysis of agnostically defined clusters was performed. Based on the phenotypes presented in figure 4 and the level of expression of each marker, gates were drawn by eye around all positive cells if the cluster had low expression of a marker on the heatmap, but only around high positive cells if the marker had high expression.

Using these definitions of Th and Treg subsets and a standard statistical approach, manual gating was able to identify a clear imbalance between pro-inflammatory Th subsets and anti-inflammatory subsets in RA. For example, out of the unstimulated dataset, manual gating was able to identify that subset 11 (CD4^+^ Tcm FoxP3^+^ IL-2^+^), a Treg subset, is decreased in RA (P = 0.0004). The same population could also be identified in the stimulated dataset (subset 17, CD4^+^ Tcm FoxP3^+^ IL-2^+^, P = 0.0002). Pro-inflammatory Th cells were also clearly identified and statistically significantly increased in RA (CD4^+^ Tcm Tbet^+^ IL-17^+^, subset 20, P = 0.0172; and subset 16, P = 0.0022; figure 8). Therefore, innovative approaches easily capture known CD4^+^ T cell subset imbalances in RA blood, even in a small sample size, by allowing for unconventional marker combinations.

**Figure 8.**
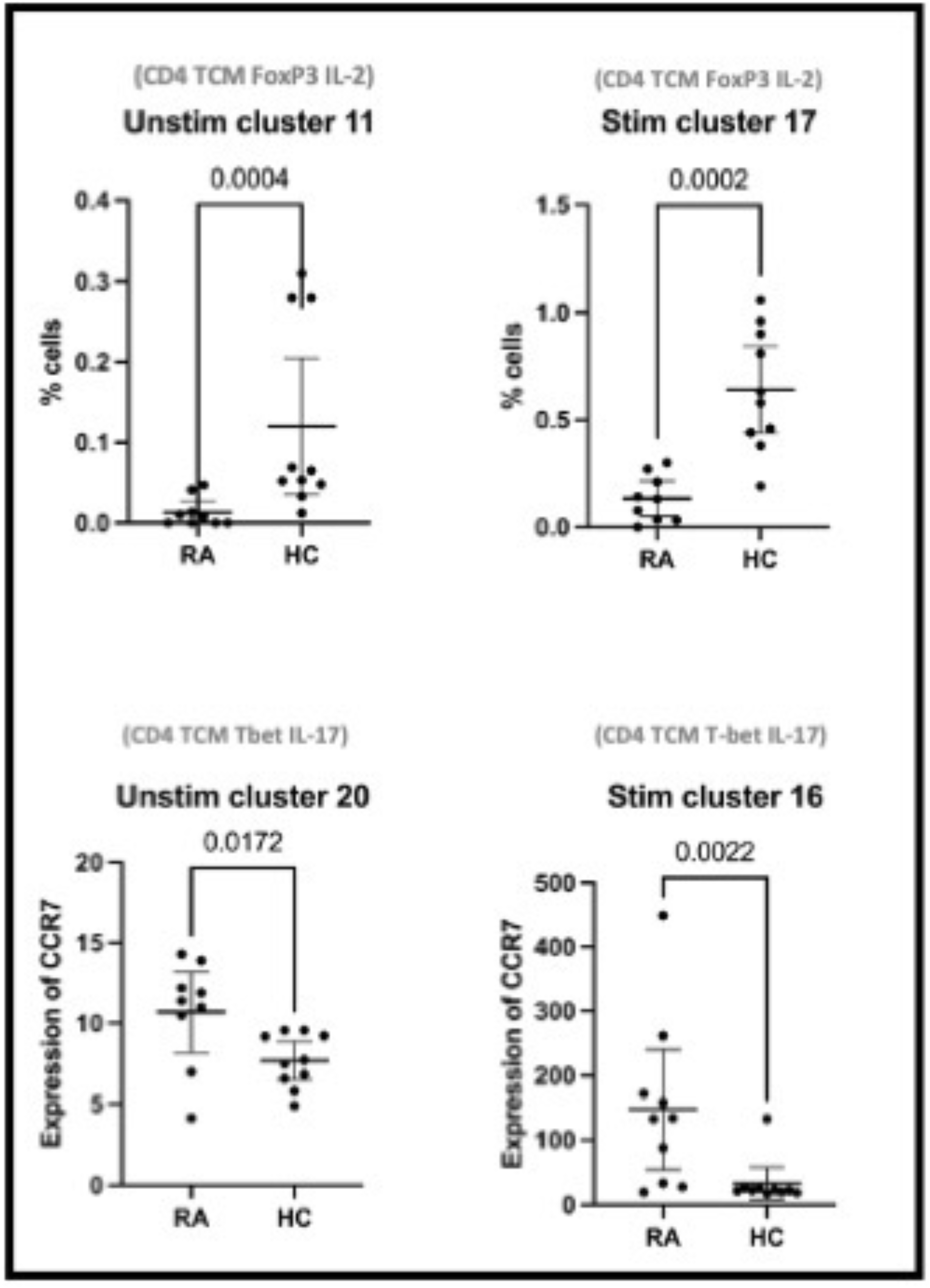
Clusters 11 and 17 represent the abundance of manually gated subsets previously identified by the CyTOF automated clustering pipeline, shown here with mean and 95% confidence intervals. Clusters 20 and 16 represent the expression of CCR7 on Th1-like IL-17^+^ cells. Points represent RA and HC samples. Mann Whitney *U P* values are shown above the pairwise comparisons. TEM: T effector memory cell. TCM: T central memory cell.

## Discussion

We show the superiority of innovative immunophenotyping and automated analytical techniques over hypothesis-driven conventional biaxial gating (based on canonical biomarkers) to identify a Th1, Th17/Th2, Treg imbalance in RA peripheral blood in small sample sizes. Our strategy allows for unusual combinations of T cell markers, for instance T-bet and IL-17, capturing the plasticity of T helper cell populations, a more difficult task to achieve with standard gating strategies.

Th17 cells are known to be phenotypically unstable ([33]). In our study, T-bet^+^ IL-17^+^ T cells are increased in RA blood. These cells have been termed Th1-like Th17 cells in the literature, co-produce IL-17 and IFNγ, and are known to contribute to inflammation in the context of autoimmunity [33–35]. Interestingly, IFNγ production in Th1-like Th17 cells has been shown to be repressed by miR-146a [35] and loss of function polymorphisms in the microRNA-146a (miR-146a) gene have been associated with an increased risk of developing RA and lupus in both European and Asian genetic association studies [36,37]; taken together, these data provide some mechanistic evidence that Th1-like Th17 cells are involved in the pathogenesis of RA. The methods outlined in this paper easily detect an increase of this subset in RA.

We also found a significant decrease in the number of IL-2^+^ FoxP3 T cells (Tregs) in RA samples compared to controls. It is well-established that IL-2 is critical in maintaining Treg function [38] and that some Treg produce IL-2 [39]. However, the relative role of paracrine versus autocrine IL-2 secretion from nearby effector T cells [40] versus IL-2^+^ FoxP3 T cells [39] is unclear. Our data suggests that IL-2-producing Tregs rather than all Treg are playing an important role in RA. Again, our methods of automated analysis were able to easily identify this non-canonical cell type, without any previous hypothesis on its association.

Due to our small sample size, our association results could theoretically represent false positive due to a sampling bias or caused by multiple testing. However, this is very unlikely, as first we confirm a plausible and previously reported imbalance of Th/Treg subsets in RA; second the mechanistic evidence supports our findings, and finally all statistical tests have been stringently corrected for multiple testing. These elements reinforce the credibility of our main conclusion: the superiority of hypothesis-free clustering of many markers in small sample sizes over standard hypothesis-driven bi-axial gating.

The hallmark of precision medicine and the primary aim of stratified medicine initiatives in RA [41] consists of stratifying patients into treatment response categories. Our study shows large heterogeneity across patients with some with a predominantly Th1 profile, others a Th17 profile, or a Th1-like Th17 profile, while others are mainly characterised by a low IL-2^+^ Treg profile. As an example, stratifying patients by their IL-2^+^ Treg titres may dictate response to low-dose IL-2 therapy, as has now been successfully trialled in lupus [42] and RA [43]. Furthermore, a Japanese group found that stratified medicine for psoriatic arthritis patients increased response rates significantly [16]. Immunophenotyped patients received ustekinumab if Th1-dominant and secukinumab if either Th17-dominant or with a Th1/Th17-hybrid phenotype resulting in clinically significant improvement in response rates [16]. The same group have good one year follow-up data [44], suggesting that the immunophenotype could be the main predictor of drug response in precision medicine. Stratifying RA patients by their Th1/Th17 phenotype, or by the Th1-like Th17 phenotype highlighted in this study, may suggest that despite the fact that previous trials of secukinumab in RA failed to reach their primary endpoint, it could be a viable treatment option in RA patient subgroups; further work is required to confirm this hypothesis. [45].

We have shown that non-biased automated analysis of large immune datasets is successful in identifying Th cell imbalance in RA with implications for precision medicine. We now plan to expand on these findings by testing their role in patient stratification for treatment response studies.

### Key messages

#### What is already known about this subject?

- Large cytometric datasets are difficult to analyse manually due to the multidimensional nature of the data.
- Manual data analysis may introduce significant bias and be underpowered to interrogate multidimensional data.

#### What does this study add?

- This study has shown that an unbiased automated clustering algorithm can successfully interrogate large cytometric datasets, finding differential expression of 2 rare T cell populations in RA patients (decreased IL-2^+^ Treg cells; increased Th1-like Th17 cells).

#### How might this impact on clinical practice?

- Understanding the underlying immunopathology of autoimmune disease is a prerequisite to finding new drug targets for treatment.
- As our knowledge of T cell heterogeneity expands, discovering rare T cell populations will directly feed into the concept of precision medicine, to: ‘treat arthritis, right first time’.

## Data Availability

All data produced are available online at https://flowrepository.org/ (repository ID FR-FCM-Z5RM).

## Contributorship

SR, RS and AB conceived the project, guided it through its stages of development, ensured progress, and contributed to the final manuscript. BM ran the laboratory experiments with help from SKH. MS pre-processed the raw mass cytometry data. BM and LM ran the data analysis and drafted the manuscript. CF supported the automated pipeline analysis and provided R code. TH and SR provided immunology expertise. All authors contributed and proof read the final version of the manuscript.

## Acknowledgments

We would like to thank Sarah Ashton (senior study coordinator) for her invaluable support, all the staff at the University of Manchester Versus Arthritis Centre for Genetics and Genomics and at the Kellgren Centre for Rheumatology, Central Manchester University NHS Foundation Trust, UK. Most importantly, we are grateful and thankful for all the patients and volunteers who took part in this study.

## Funding and grant awards

This research was supported by Versus Arthritis (grant 21754), the NIHR Manchester Biomedical Research Centre, and the Wellcome Trust Institutional Strategic Support Fund (WT ISSF) in Precision Medicine and Single Cell Research. AB is an NIHR Senior Investigator. BM is an NIHR-funded Academic Clinical Fellow.

