## Supplementary Methods for "CD4^+^ T cell subset imbalances in rheumatoid arthritis"

Supplementary material

Supplementary methods

Stimulated and unstimulated CD3^+^ T cells were incubated in 2.5 µM cisplatin (Fluidigm, Cambridge, UK) plus 1:100 Human TruStain FcX Fc receptor blocker (BioLegend) in PBS for 3 minutes at room temperature, followed by quenching with Maxpar Cell Staining Buffer (CSB; Fluidigm) twice. An extracellular antibody stock was made using 0.5 µl of antibody per test in 50 µl of CSB, added to each sample and incubated for 30 minutes at room temperature. Cells were then washed twice with CSB and fixed in eBioscience Foxp3 / Transcription Factor Staining Buffer Set (Fisher Scientific UK Ltd) for 60 minutes on ice, made up as per the manufacturer’s instructions. Cells were permeabilised with 10 % eBioscience permeabilisation buffer (Fisher Scientific UK Ltd) and stained for intracellular antigens by making up an antibody cocktail of 0.5 µl in 100 µl permeabilisation buffer per test for 30 minutes in the dark on ice. A 5 µM solution of Maxpar Cell-ID Intercalator-Ir (Fluidigm) was made up in Maxpar Fix & Perm Buffer (Fluidigm) and 200 µl was added to each well and kept at 4 °C overnight, or until the samples could be run on the mass cytometer. Immediately prior to running samples, cells were washed twice with ultrapure water from a Milli-Q water system (Millipore, Merck). Maxpar EQ Four Element Calibration Beads (Fluidigm) were made up in ultrapure water at a ratio of 1:10 (4EQ buffer) and used to resuspend the cells to attain a concentration of 700,000 cells/ml wherever possible. The sample was then passed through a 40 µm Corning Falcon test tube with cell strainer snap cap (Fisher Scientific UK Ltd), prior to running on the mass cytometer. Samples were run on two Helios mass cytometers (Fluidigm) by the Longwood Medical Area CyTOF Core at the Dana-Farber Cancer Institute, Boston, USA.

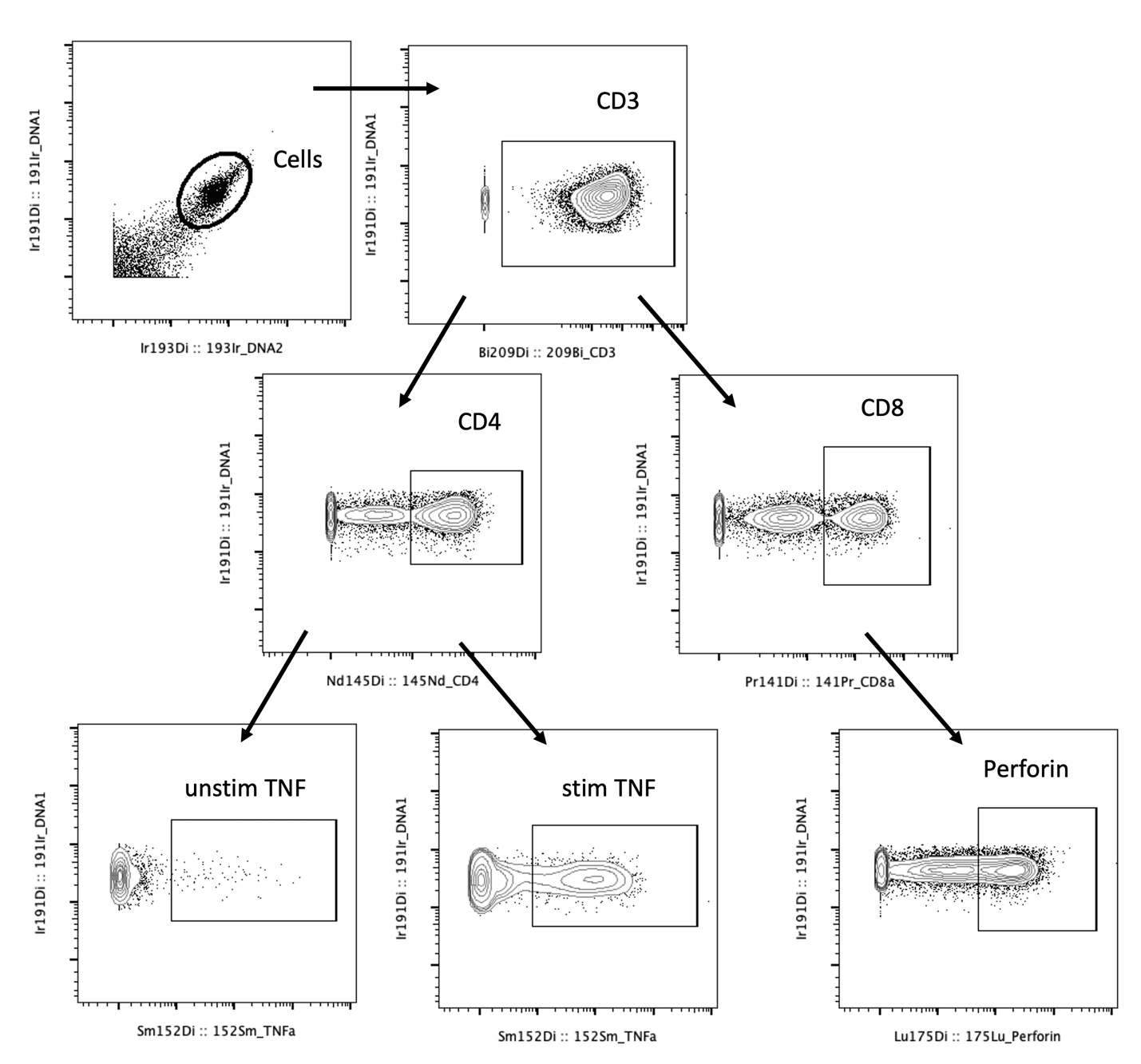

**Supplementary figure 1.** Representative dot plots showing manual gating of T cell subsets. Biaxial cytometry dot plots showing CD4^+^ and CD8^+^ T cell gating, following by unstimulated and stimulated TNF in CD4^+^ and perforin in CD8^+^ T cells.

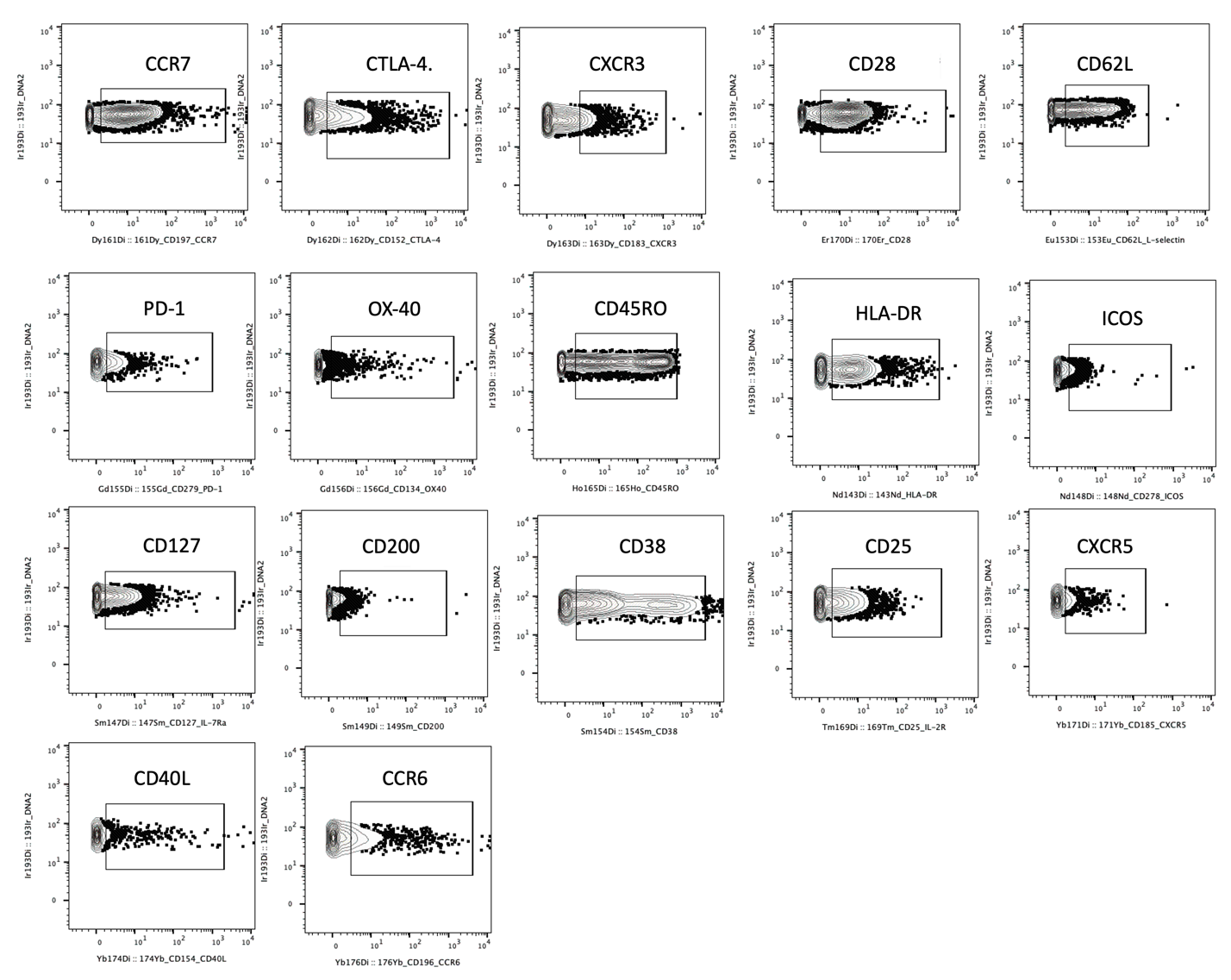

**Supplementary figure 2A.** Manual gating of cell surface markers to determine lower and upper cut off points. All dot plots are of one sample but are representative of the entire dataset.

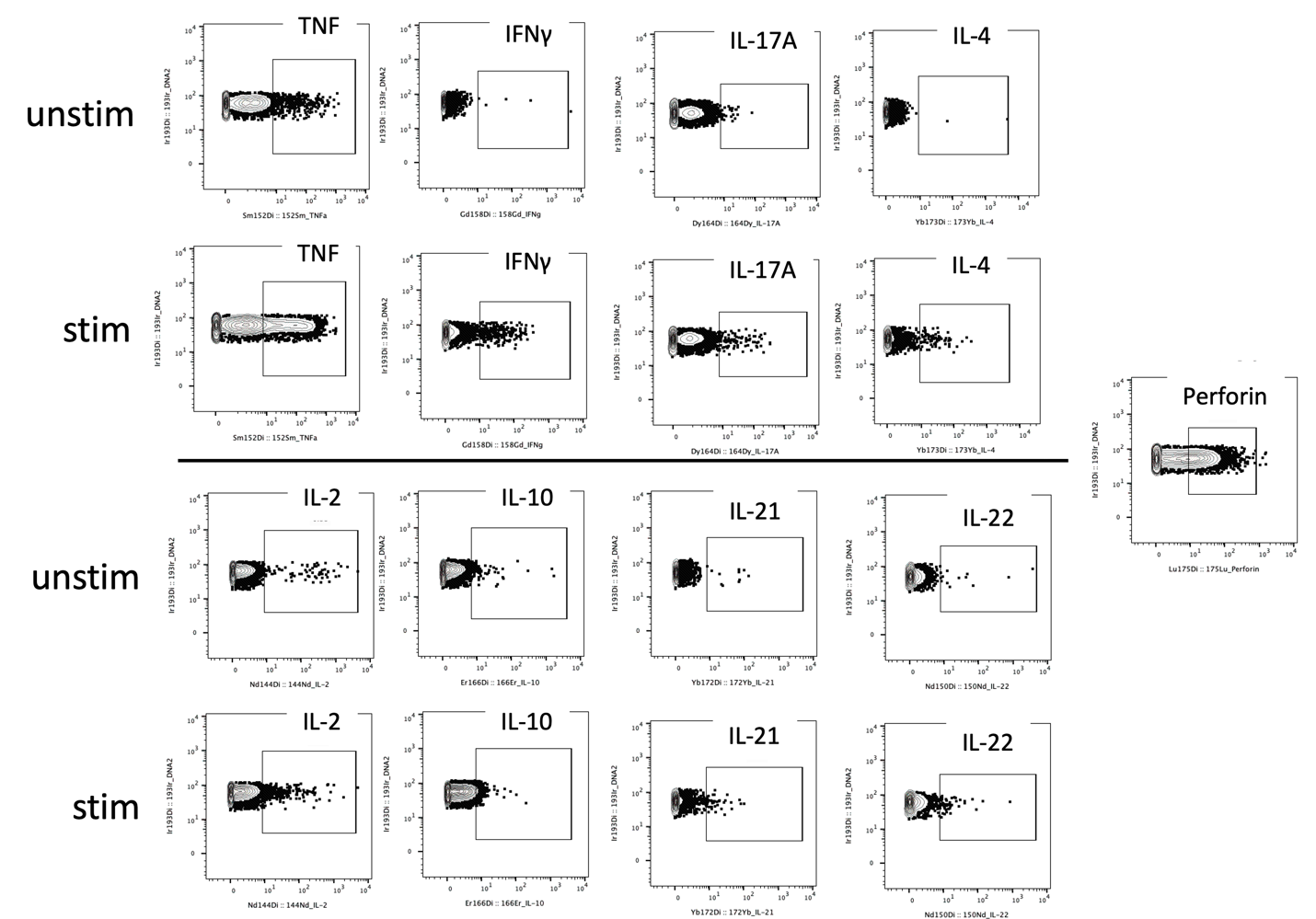

**Supplementary figure 2B.** Manual gating of intracellular markers to determine lower and upper cut off points. Both unstimulated and stimulated samples are shown to represent cytokine expression. All dot plots are of one sample but are representative of the entire dataset.

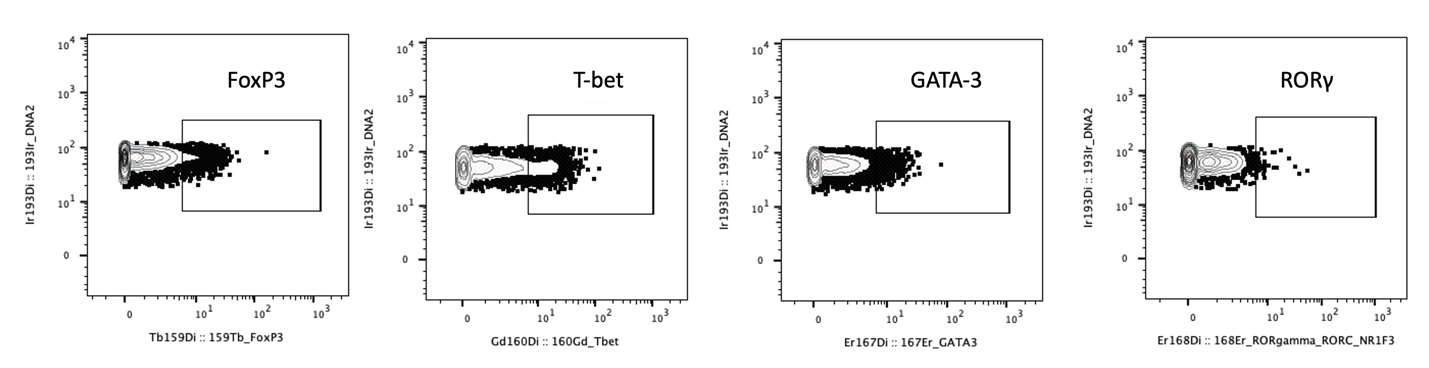

**Supplementary figure 2C.** Manual gating of transcription factors to determine lower and upper cut off points. All dot plots are of one sample but are representative of the entire dataset.

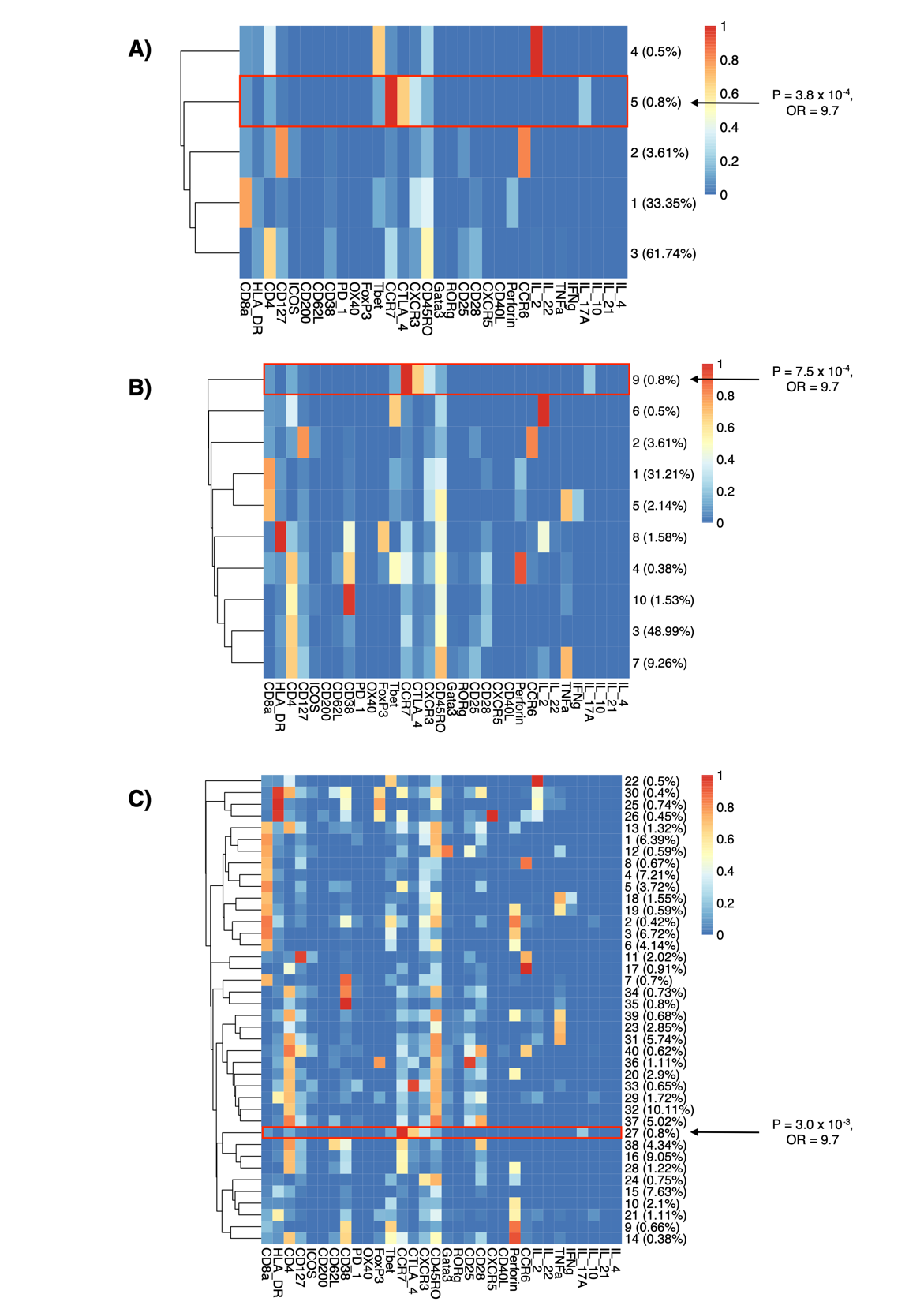

**Supplementary figure 3.** Automated clustering of stimulated T cell samples (*n* = 10 RA, 10 healthy controls) into **A)** 5, **B)** 10 and **C)** 40 clusters. This cluster titration was performed to evaluate the most appropriate cluster number and the influence of changing this parameter on the consistency of significant findings. We concluded that there was over-clustering present with 40 clusters. Five and 10 clusters did not identify subsets with sufficient granularity. Red boxes highlight the presence of CD4+ IL-17+ T-bet+ cells in each analysis with a consistent finding of differential abundance of this subset in RA vs healthy samples, supporting the robustness of the findings. *P* values and ORs were derived from MASC.
